# SOX9 Expression in Colorectal Adenomas Improves Surveillance Colonoscopy Risk Stratification in a Bowel Screening Population

**DOI:** 10.1101/2024.06.28.24309576

**Authors:** Sara Samir Foad Al-Badran, Christopher Bigley, Mark Johnstone, Aula Ammar, Alexander Winton, Jennifer Hay, Jean Quinn, Jakub Jawny, Ditte Andersen, Natalie Fisher, Philip D. Dunne, Noori Maka, Gerard Lynch, Stephen McSorley, Joanne Edwards INCISE Collaborative

## Abstract

**Objectives:** Adenomas are known precursors to colorectal cancer (CRC). Current UK post-polypectomy surveillance guidelines use polyp size, numbers, and histology to stratify the risk of patients developing metachronous polyps or CRC. However, these risk guidelines suffer from poor predictive value, often leading to under/over surveillance.

**Design:** Adenomas removed from 1257 patients at bowel screening colonoscopy were retrospectively identified to investigate mutational profile and protein expression trends associated with the detection of metachronous polyps or CRC. The presence or absence of metachronous polyps or CRC was recorded 6 months to 6 years after index polypectomy.

**Results:** *APC* and *KRAS* were the most mutated genes in these patients (87% and 34% respectively), and both were significantly co-occurring with the 6th most mutated gene *SOX9* (17% co-occurring with *APC*, p=0.047; 23% co-occurring with *KRAS*, p=0.012). High SOX9 cytoplasmic expression was significantly associated with the detection of metachronous polyps or CRC (HR 1.543, p=0.001) and improved high risk stratification when combined with BSG2020 guidelines versus guidelines alone (HR 2.626, p<0.0001). High cytoplasmic SOX9 alone and in combination with current guidelines was an independent predictor of metachronous polyps or CRC according to various regression models. This was validated in an independent test dataset, where high cytoplasmic expression was significantly associated with the detection of metachronous polyps or CRC (HR 1.654, p=0.012) and enhanced risk stratification when combined with BSG2020 guidelines versus guidelines alone (HR 2.473, p=0.0018).

**Conclusion:** High cytoplasmic SOX9 expression within adenomas is associated with shorter time to detection of metachronous polyps or CRC.

## INTRODUCTION

Colorectal cancer (CRC) is ranked third in diagnosis and second in cancer deaths globally[1–4]. The first step in the development of CRC is the appearance of precursor polyps in the bowel[5]. The two most common types of premalignant polyps are adenomatous polyps (adenomas), from which ∼85% of CRCs develop, and serrated polyps[6–8]. Adenomas develop through the well-established adenoma-carcinoma sequence, while serrated polyps develop via the serrated polyp pathway[9,10].

The transformation of benign polyps to malignancy is a process estimated to take between 7-15 years, presenting a window of opportunity for early detection and intervention[11]. As a result, bowel screening programs (BSPs) have been established worldwide to both diagnose early asymptomatic cancer and identify and remove premalignant polyps[12]. Premalignant polyps (removed by polypectomy) are detected in 25-50% of individuals aged 50-74 years screened through the Scottish BSP. It is estimated that 50% of the patients will develop future/metachronous polyps or cancer, making further surveillance colonoscopies necessary[8,11,13].

The current British Society of Gastroenterology (BSG) guidelines for post-polypectomy surveillance stratifies patient risk of future metachronous polyps based on the size, number, and degree of cellular atypia at the first (index) polypectomy. Patients classified as high risk undergo a surveillance colonoscopy after three years[8], but only a small proportion go on to develop metachronous polyps with advanced features or cancer[14]. For these patients there is limited benefit in terms of early cancer detection when adhering to the current surveillance framework. Of more notable concern are the number of cases classified as low risk by these parameters who go on to develop metachronous polyps.

Several genomic and immunohistochemical markers associated with metachronous polyp risk have been previously identified within the literature[15]. INtegrated teChnologies for Improved polyp SurveillancE (INCISE) is a large, retrospective, multi-partner collaborative project which aims to better stratify future polyp risk through detailed analysis of adenoma tissue beyond their phenotypic histopathological features. It is hoped that such an approach could be used to refine surveillance protocols, reducing unnecessary surveillance and the burden on stretched endoscopy services, whilst increasing the detection of high risk metachronous polyps. In this paper, we aim to examine adenomas removed at bowel screening colonoscopies regarding their mutational landscape, gene expression, and protein expression in relation to the likelihood of detection of metachronous polyps or CRC at surveillance to determine whether such characteristics can improve on the existing risk categories.

This study presents a mutational and protein expression analysis of SOX9 in pre-cancerous index human colorectal polyps. For the first time, it proposes a new method to improve the current BSG2020 Guidelines based in combination with protein expression data.

## MATERIALS AND METHODS

### Patient Cohort

This study used human tissue collected for the **In**tegrated te**C**hnologies for **I**mproved polyp **S**urveillanc**E** (INCISE) collaborative from polypectomies performed within the Scottish BSP in Greater Glasgow and Clyde between 2009-2016 in patients who underwent further colonoscopy between 6 months and 6 years after the index procedure[15]. The pathology of the excised index polyp specimens was outlined in local histopathology reports. Characteristics including the number of polyps present, histology (adenoma or serrated), morphology (presence or absence of villous features), location (rectum, left or right side of the colon), size (<10mm or ≥10mm), presence of advanced polyps were recorded and BSG2020 Guidelines categorised. The presence or absence of metachronous polyps or CRC at surveillance was determined using electronic endoscopy reporting software (Ver. 2.5, Unisoft GI Reporting Software, Unisoft Medical Systems, UK) and electronic pathology database, TelePath. This project received ethical approval from the West of Scotland Research Ethics Committee (22/WS/0020) and patient information is held within Safe Haven. There was no patient involvement in the design of this study.

### Sample DNA Sequencing

Sample sequencing and variant calling was performed by the Genomic Innovation Alliance (Glasgow, UK) using the Agilent SureSelect XT2 HS2 method (Agilent, Santa Clara, CA). Regions of interest were enriched with the Agilent SureSelect CancerPlus panel (Design ID: S3225252, included genes listed in S-Table 3) and the quality and quantity of libraries determined by TapeStation using a D1000 ScreenTape (5067-5582, Agilent, Santa Clara, CA).

### Mutational Analysis

Mutational analysis was performed on Rstudio[16] (2023.12.0+369; MA, USA) using the Maftools[17] (Ver. 2.18.0) package under the BiocManager Repository[18] (Ver. 1.30.22). Oncoplots were generated using the “oncoplot” function. Pairwise Fisher’s Exact test was performed to establish co-occurring and mutually exclusive mutated gene pairs using the “somaticInteractions” function. Lollipop plots were generated using the MutationMapper tool on cBioPortal[19–21]. All statistical significance for mutational analysis was set at an adjusted p-value (PAdj) of <0.05.

### Immunohistochemistry and Visualization

Immunohistochemistry (IHC) was employed to detect the protein expression of SOX9 in adenoma tissue microarrays (TMA). Anti-SOX9 (Cat. Ab5335, Millipore, Gillingham, UK) was validated for specificity and sensitivity (S-Fig.2) as described in the supplementary methods. TMAs were then stained at a concentration of 1:4000 using the Leica Bond RX autostainer, with the antigen retrieval step using a pH9 buffer for 20 minutes. Matched isotype and no-antibody negative controls were included, as well as breast tissue positive controls (Fig.2C). The tissue was scanned using the Nanozoomer Scanner (Hamamatsu Photonics, Hertfordshire, UK), and visualized on NZConnect (Ver. 1.1.0, Hamamatsu Photonics, Hertfordshire, UK).

### Protein Expression Assessment

Protein expression was assessed digitally using QuPath Digital Pathology Platform[22] to determine the weighted histoscore (H-Score) for SOX9, as previously described[23]. In brief, slides were dearrayed and stain vectors were estimated during pre-processing to improve colour deconvolution quality. Then, cell objects were detected using watershed cell detection, followed by annotation of different tissue types (epithelium and lamina propria). A random trees classifier (Fig.2C) was trained using a variety of features on an independent set of sections and applied to detected cell objects to determine cell identity. Three intensity thresholds were set on the DAB colour deconvolved layer for each cellular compartment expressing SOX9 to represent negative, weak, moderate, and strong staining (Fig.2C), providing H-Scores between 0-300. H-Scores generated by QuPath were compared to manual assessment to ensure consistency. Both cores representing the luminal epithelium (LE) for the same patient were averaged to find a mean score, as were the two representing the basal epithelium (BE; S-Fig.1). Examples of low and high expression SOX9 in each expressing compartment are represented in Fig.2C.

### Statistical Analysis

IHC H-Scores from the training dataset were imported into Rstudio to generate thresholds for expression dichotomization using the *Survminer*[24] (Ver.0.4.9) and *Maxstat*[25] (Ver. 0.7-25) packages. The suggested cut-points were 216.06 for nuclear SOX9 and 177.39 for cytoplasmic SOX9 (S-Fig.4). However, these are not clinically usable values as they are difficult to ascertain visually by pathologists. Hence, we chose a close approximation of 215 as a threshold for nuclear SOX9 and 200 for cytoplasmic SOX9. These thresholds were applied to the test dataset.

1-survival analysis with log rank statistics and 95% Confidence Intervals (95% CIs) was carried out to generate Kaplan-Meier Curves with detection of metachronous polyp or CRC set as an endpoint, using *Survminer* and *Survival*[26,27] (Ver. 3.5-7). SPSS (Ver. 28.0.1.1 (15), IBM, NY, USA) was used for to assess associations between SOX9 expression and clinicopathological characteristics using *X*^2^ tests where p<0.05 was considered statistically significant. Univariate Cox regression survival analysis was used to determine hazard ratios (HRs) and 95% confidence intervals (CIs). Multivariable Cox regression survival analysis using a backward conditional elimination model and a statistical significance threshold of p<0.05, was performed to identify independent predictors of metachronous polyps or CRC.

### RNA Sequencing

TempO-Seq™ (Biospyder Technologies, Carlsbad, CA, USA) whole transcriptome profiling was performed on 816 patients from the INCISE cohort, according to the manufacturer’s instructions using whole FFPE tissue sections.

### Transcriptomic Processing and Analysis

Quality control and preprocessing was carried out in RStudio (R 4.2.3). Probe variance across all samples was calculated using the *sapply* function. Probes with a variance less than the 25th percentile were removed (n=5640 probes). Samples with a low read count (< 95% CI [<2.28×106 counts]) were identified (*stats* Ver. 4.2.3) and removed (n=110), as well as samples which did not match batch information (n=5). A subset of patients with matched IHC data was selected for this study (n=816). The counts were batch corrected using *ComBat_seq* (sva Ver. 3.46.0). Probe IDs were mapped to gene symbols with duplicated genes collapsed using *MaxMeans*[28,29] (*WGCNA* Ver. 1.72-1), resulting in 14,993 genes. Normalised counts were generated using quantile normalisation and log2 +1 transformed.

All the transcriptome analysis was performed using R (4.3.3) in RStudio. *DESeq2* (Ver. 1.42.1) was used to perform differential gene expression analysis, and any results were plotted using *ggplot2* (Ver. 3.5.0).

## RESULTS

### Mutational Patient Cohort Description

Tissue from 598 patients was sequenced for mutational analysis. 446 (75%) were from males and 570 (95%) of patients were of screening age. 421 (71%) of specimens were from the left colon, and 504 (84%) were without high grade dysplastic features. 412 (69%) of patients presented with >1 index polyp, and 356 (60%) of polyps had tubulovillous/villous architecture (Fig.1A).

**Figure 1.**
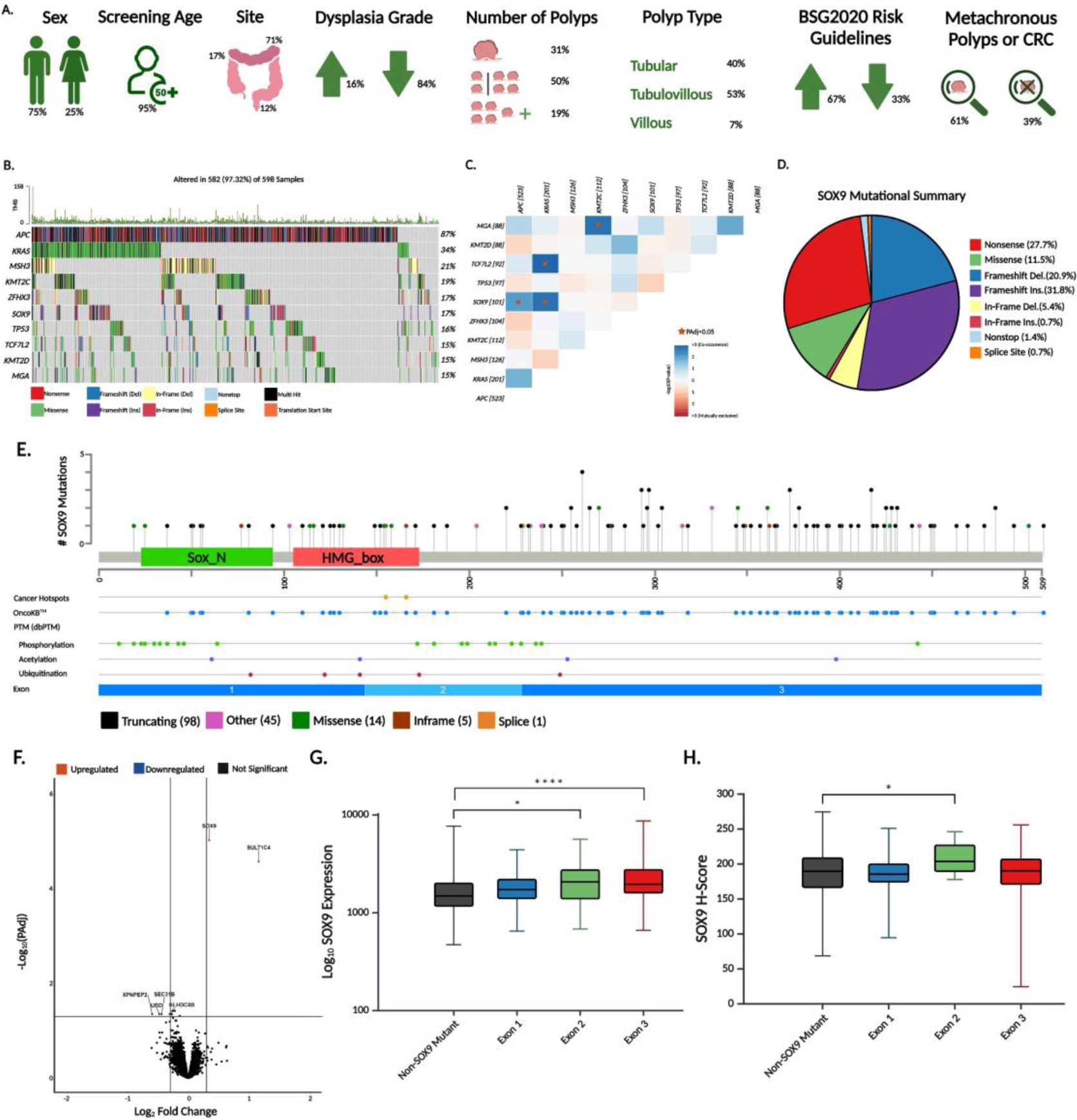
Mutational Landscape Investigation of INCISE Cohort. **[A]** Description of mutational cohort in relation to clinical features. **[B]** Oncoplot of INCISE patient samples depicting the top 10 mutated genes, ranked by percentage of samples affected for each gene, and then alphabetically by gene name. **[C]** Co-occurrence analysis of mutations between genes. Stars indicate significant co-occurrence with PAdj<0.05. Number of cases affected is shown in square brackets. **[D]** Mutational summary of *SOX9*. **[E]** Lollipop plots of *SOX9* amino acid changes across the length of the gene. **[F]** Volcano plot of differentially expressed genes. **[G]** Box plot of the logged mRNA expression of *SOX9* against patients with *SOX9* mutations grouped by exonic location, and non-*SOX9* mutant patients. Asterisks denote significance (p<0.05) by the Mann-Whitney U Test. **[H]** Box plot of the protein expression of SOX9 represented by a H-Score against patients with *SOX9* mutations grouped by exonic location, and non-*SOX9* mutant patients. Asterisks denote significance (p<0.05) by the Mann-Whitney U Test.

### Mutational Landscape of Index Polyps

From each patient, the largest most advanced polyp was mutationally profiled. 97% of sequenced samples were mutated, with 87% and 34% exhibiting *APC* and *KRAS* mutations, respectively. The top 10 most commonly mutated genes are shown in Figure 1B. Somatic interactions analysis (Fig.1C, S-Table 4) revealed the most significantly co-occurring mutated gene pairs to be *TCF7L2* and *KRAS* (PAdj=0.009, event ratio 22%), and *MGA* and *KMT2C* (PAdj=0.012, event ratio 19%). *SOX9* mutations were significantly co-occurring with mutations in both *KRAS* (PAdj=0.012, event ratio 23%), and *APC* (PAdj=0.047, event ratio 22%). No two genes were found to be significantly mutually exclusive. Although SOX9 was included in the 3^rd^ and 4^th^ most mutated pairs, its co-occurrence with the top two mutated genes, *APC* and *KRAS,* is noteworthy. This, along with its involvement in WNT signaling led to us pursuing it for further analysis.

*APC* and *KRAS* were the two most mutated genes, in agreement with the literature on cancer studies. Co-occurrence of *SOX9* mutation with *APC and KRAS* in adenomas has not previously been reported. As seen in Figure 1D, *SOX9* mutations were mostly frameshift insertions or nonsense mutations (31.8% and 27.7%, respectively). Frameshift deletions made up 20.9% of the mutational burden of *SOX9*, while missense and in-frame deletions made up 11.5% and 5.4% respectively.

60% of the mapped mutations appeared to be truncating mutations. OncoKB curated alterations of SOX9 are almost always loss-of-function and considered “likely oncogenic”. Exons 1 and 2 had most of the post-translational modifications, the most common being phosphorylation. However, Exon 3 had the highest mutational burden (Fig.1E).

### SOX9 Mutations are Associated with Differential SOX9 Gene and Protein Expression in Adenomas

Comparison of differentially expressed genes dependent on SOX9 mutational status revealed that *SOX9* (PAdj<0.0001, Log2 Fold Change 0.35) mRNA expression was upregulated in *SOX9* mutated cases suggesting that *SOX9* mutations have a functional impact (Fig.1F). In addition, *SULT1C4* (PAdj<0.0001, Log2 Fold Change 1.2) was upregulated and four genes were downregulated, *XPNPEP2* (PAdj=0.04, Log2 Fold Change −0.6)*, UBD* (PAdj=0.04, Log2 Fold Change −0.5)*, SEC31B* (PAdj=0.04, Log2 Fold Change −0.4), and *KLHDC8B* (PAdj=0.04, Log2 Fold Change −0.3). As *SOX9* was observed to accumulate mutations along its amino acid structure (Fig.1E), we explored whether the gene expression was associated with mutations classified by exonic location (Fig.1G). When compared with patients that did not have *SOX9* mutations, mutations in exon 2 were significantly associated with upregulation of *SOX9* gene (p=0.02) and protein expression (p=0.03), whilst mutations in exon 3 were significantly associated with upregulated *SOX9* gene expression (p<0.0001) but not protein expression (Fig.1H).

### TMA Cohort Description

The INCISE TMA Cohort was comprised of a training dataset which included two thirds of the patients (n=868), and a test dataset which included the remaining third (n=389). Due to low numbers, polyps smaller than 10mm and all non-adenomas (serrated/hyperplastic/etc.) were excluded from analysis (Fig.2A). Unless otherwise specified, all analysis was performed independently for the training and test datasets. Frequencies for clinicopathological variables were similar between training and test datasets. In the training set, 538 (66%) of patients were stratified as high risk according to current BSG2020 Guidelines, of which 61% had a metachronous polyp or CRC, whilst 34% were stratified as low risk, of which 45% had a metachronous polyp or CRC. (Fig.2B; Table 1; S-Table 1).

**Figure 2.**
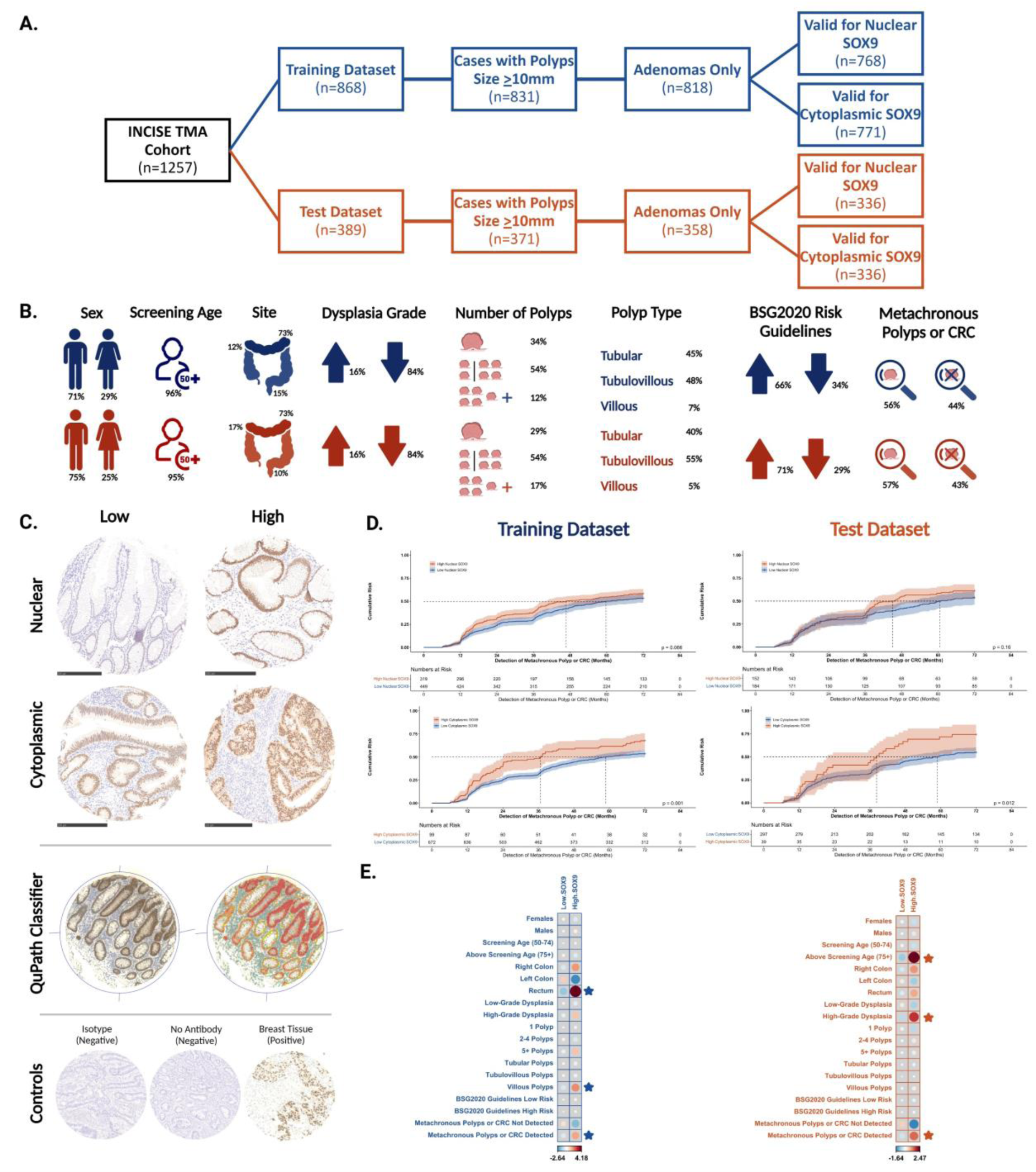
Investigation of SOX9 as a Predictive Biomarker for the Development of Metachronous Polyps or CRC. **[A]** CONSORT Diagram of INCISE TMA Cohort into separate (blue) training and (orange) test datasets. **[B]** Description of clinical characteristics in the training (blue) and test (orange) patient datasets. **[C]** Staining examples of low and high expression of SOX9 in the nucleus and cytoplasm of polyp tissue (top). QuPath classifier for tissue type and staining intensity where red is strong epithelium, orange is moderate epithelium, yellow is weak epithelium, blue is negative epithelium and green is lamina propria (middle). Negative and positive *in-assay* controls (bottom). **[D]** 1-Survial analysis of (top) nuclear and (bottom) cytoplasmic SOX9 in the (left) training and (right) test datasets. Shading is indicative of 95% CI. Dotted lines are time to median risk. **[E]** Corrplots of associations between low and high cytoplasmic SOX9 expression and clinical characteristics in the (blue) training and (orange) test datasets. Dot size and color intensity (as directed by the z-score indicator) suggests a high correlation between variables. Stars denote statistically significant associations.

**Table 1.**
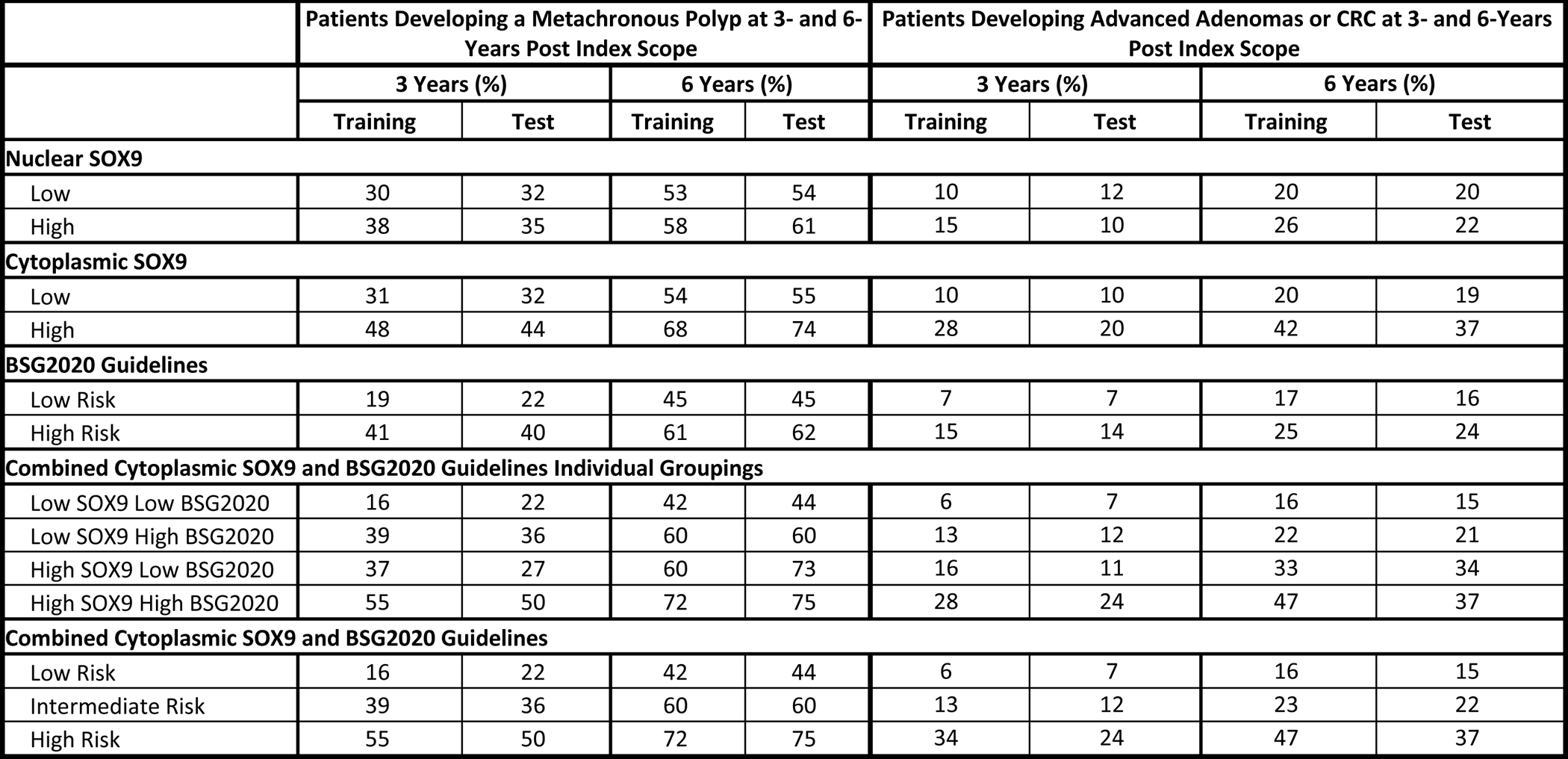
Cumulative Proportion of Patients Developing Metachronous Polyps and Developing Advanced Adenomas or CRC at 3- and 6-Years Post Index Scope.

### Cytoplasmic SOX9 Expression is Associated with Metachronous Polyp or CRC Detection

SOX9 was observed in the nucleus and cytoplasm of epithelial adenoma cells and in cores of luminal and basal epithelium (S-Fig.1, Fig.2C). H-Scores from each epithelium region were combined to calculate total adenoma epithelium protein expression for each adenoma(S-Fig.3). After dichotomizing H-Scores into low SOX9 and high SOX9 for each cellular compartment (thresholds of 215 for nuclear and 200 for cytoplasmic), 1-survival analysis (Fig.2D) on the training dataset revealed that nuclear SOX9 protein expression was not statistically associated with the detection of metachronous polyps or CRC. Cytoplasmic SOX9 was significantly associated (p=0.001) with time to detection of metachronous polyps in the training (p=0.001) and test (p=0.012) datasets, with 48% of those with high cytoplasmic SOX9 having a metachronous polyp at 3 years compared to 31% with low SOX9 expression (Table 1).

### Cytoplasmic SOX9 Expression in Adenomas is Associated with Adverse Pathological Features

In the training dataset, high cytoplasmic SOX9 was positively associated with rectal polyps (p<0.001), and villous features (p=0.009), and the detection of metachronous polyps or CRC (p=0.008). In the test dataset positive associations were observed between high cytoplasmic SOX9 and high-grade dysplasia (p=0.034; Fig.2E, S-Table 2).

### Combining SOX9 with BSG2020 Guidelines Improves Risk Stratification

By applying the current BSG2020 Guidelines to our cohort, there was a statistically significant split (p<0.0001) between low risk and high risk patients in the training dataset (Fig.3A). 41% and 61% of the high risk group had a metachronous polyp 3 years (first follow-up scope) and 6 years (second follow-up scope) after the index scope, respectively. This was validated in the test dataset (p=0.0015; Fig.3B), where 40% and 62% of the high risk group had a metachronous polyp 3 and 6 years after the index scope (Table 1).

**Figure 3.**
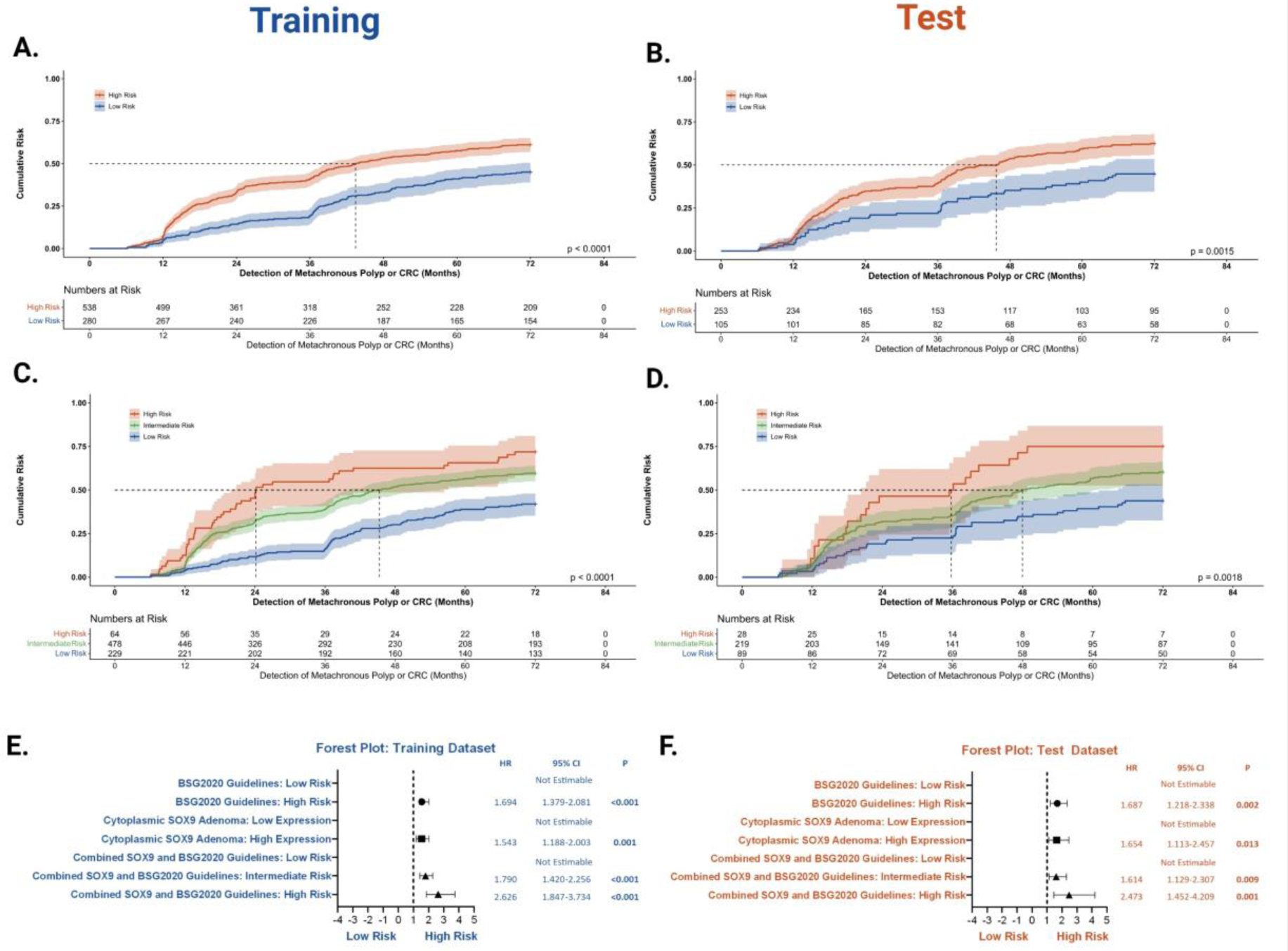
Combination of Cytoplasmic SOX9 with BSG2020 Guidelines for Improved Risk Stratification. 1-Survival analysis of BSG2020 guidelines in the **[A]** training (p<0.0001) and **[B]** test (p=0.0015) datasets with patients split into low and high risk. 1-Survival analysis of risk stratification based on cytoplasmic SOX9 combined with BSG2020 Guidelines in the **[C]** training (p<0.0001) and **[D]** test (p=0.0018) datasets. Dotted lines are time to median risk (low risk patients do not make it to median risk and so do not have a line). Forest plots cytoplasmic SOX9 expression effect on the detection of metachronous polyps or CRC and BSG2020 guidelines, alone and in combination in the **[E]** training and **[F]** test datasets with HRs and 95% CIs.

When SOX9 expression was added to these criteria, 4 groups were generated, “Both Low” for those who have low SOX9 expression and are BSG2020 guidelines low risk, “Both High” for those who have high SOX9 expression and are BSG2020 guidelines high risk, and two intermediate groups of “High SOX9 Low BSG2020”, and “Low SOX9 High BSG2020”. In the training dataset (p<0.0001; S-Fig.5A), 55% of the “Both High” group had a metachronous polyp or CRC 3 years after the index scope (Table 1), while in the test dataset (p=0.0047; S-Fig.5B), 50% of the “Both High” group had a metachronous polyp 3 years after the index scope (Table 1). The intermediate groups of “High SOX9 Low BSG2020”, and “Low SOX9 High BSG2020” had comparable proportions of patients with metachronous polyps detected 3 years after the index scope (37% and 39%, respectively), and were combined into a single group. This led to the final groupings of “Low Risk” patients, “Intermediate Risk” patients, and “High Risk” patients.

In the training dataset (Fig.3C), there was a significant difference in the proportion of patients with metachronous polyps or CRC between groups (p<0.0001), where 16% of the Low Risk group had an event 3 years after the index scope, when compared to 55% of the High Risk group (Table 1). The same was true in the test dataset (p=0.0018; Fig.3D), where 22% of the Low Risk group had a metachronous polyp detected 3 years after the index scope versus 50% for the High Risk group.

Finally, when investigating the associations between our risk stratification model and the type of metachronous lesions seen after the index scope in the training dataset, we found that only 13% of the Low Risk group had an advanced adenoma or CRC at follow-up, while 38% of the High Risk group had an advanced adenoma or CRC at follow-up. This was validated in the test dataset (S-Table 5). These results suggest an advantage to including biomarker expression to current surveillance colonoscopy risk stratification criteria.

### SOX9 and BSG2020 Guidelines Combination Is an Independent Predictive Factor of Metachronous Polyps or CRC

Univariate Cox regression was carried out on the training dataset and of the clinical features included (Table 2), male sex (HR 1.506; p<0.001), left colonic polyps (HR 0.615; p<0.001), multiple index polyps (HR 1.546 for 2-4 polyps and HR 2.549 for 5+ polyps; both p<0.001), high risk according to BSG2020 Guidelines (HR 1.694; p<0.001), high cytoplasmic SOX9 expression (HR 1.543; p=0.001), and the “Intermediate Risk” and “High Risk” groups of the SOX9 and BSG2020 Guidelines combination (HR 1.790 for Intermediate Risk and HR 2.626 for High Risk; p<0.001 for both), were all significantly associated with the detection of metachronous polyps or CRC and entered into multivariate analysis (Fig.3E).

**Table 2.** Cytoplasmic SOX9 Expression, Clinicopathological Characteristics, and the Detection of Metachronous Polyps or CRC in the INCISE Training Dataset.

|  |  | Univariate |  |  | Multivariate |  |  |  |  |  |  |  |  |
| --- | --- | --- | --- | --- | --- | --- | --- | --- | --- | --- | --- | --- | --- |
|  |  |  |  |  | Model 1:<br>BSG2020 Guidelines<br>Components |  |  | Model 2:<br>BSG2020 Guidelines |  |  | Model 3:<br>Combined Cytoplasmic SOX9 and<br>BSG2020 Guidelines |  |  |
|  |  | Hazard<br>Ratio | 95% CI | P | Hazard<br>Ratio | 95% CI | P | Hazard<br>Ratio | 95% CI | P | Hazard<br>Ratio | 95% CI | P |
| Sex | Female | 1.0 | - | - | 1.0 | - | - | 1.0 | - | - | 1.0 | - | - |
|  | Male | 1.506 | 1.214-1.868 | <0.001 | 1.446 | 1.156-1.810 | 0.001 | 1.463 | 1.169-1.830 | 0.001 | 1.463 | 1.169-1.830 | 0.001 |
| Age | Screening Age | 1.0 | - | - | - | - | - | - | - | - | - | - | - |
|  | Above Screening Age | 0.844 | 0.513-1.391 | 0.506 | - | - | - | - | - | - | - | - | - |
| Site | Right Colon | 1.0 | - | - | 1.0 | - | - | 1.0 | - | - | 1.0 | - | - |
|  | Left Colon | 0.615 | 0.473-0.800 | <0.001 | 0.656 | 0.501-0.858 | 0.002 | 0.650 | 0.497-0.850 | 0.002 | 0.655 | 0.501-0.856 | 0.002 |
|  | Rectum | 0.844 | 0.608-1.172 | 0.311 | 0.912 | 0.652-1.275 | 0.589 | 0.911 | 0.652-1.275 | 0.588 | 0.919 | 0.656-1.288 | 0.625 |
| High Grade Dysplasia | Absent | 1.0 | - | - | - | - | - | Omitted From Model |  |  | Omitted From Model |  |  |
|  | Present | 1.126 | 0.881-1.439 | 0.342 | - | - | - |  |  |  |  |  |  |
| Number of Polyps | 1 | 1.0 | - | - | 1.0 | - | - | Omitted From Model |  |  | Omitted From Model |  |  |
|  | 2-4 | 1.546 | 1.249-1.915 | <0.001 | 1.551 | 1.242-1.936 | <0.001 |  |  |  |  |  |  |
|  | 5+ | 2.549 | 1.905-3.410 | <0.001 | 2.340 | 1.724-3.177 | <0.001 |  |  |  |  |  |  |
| Polyp Type | Tubular | 1.0 | - | - | - | - | - | - | - | - | - | - | - |
|  | Tubulovillous | 0.932 | 1.219-1.127 | 0.466 | - | - | - | - | - | - | - | - | - |
|  | Villous | 0.936 | 0.638-1.372 | 0.735 | - | - | - | - | - | - | - | - | - |
| BSG2020 Guidelines | Low Risk | 1.0 | - | - | Omitted From Model |  |  | 1.0 | - | - | Omitted From Model |  |  |
|  | High Risk | 1.694 | 1.379-2.081 | <0.001 |  |  |  | 1.677 | 1.354-2.076 | <0.001 |  |  |  |
| Cytoplasmic SOX9 Adenoma | Low Expression | 1.0 | - | - | 1.0 | - | - | 1.0 | - | - | Omitted From Model |  |  |
|  | High Expression | 1.543 | 1.188-2.003 | 0.001 | 1.397 | 1.068-1.828 | 0.015 | 1.465 | 1.124-1.911 | 0.005 |  |  |  |
| Combined Cytoplasmic SOX9 and BSG2020 Guidelines | Low Risk | 1.0 | - | - | Omitted From Model |  |  | Omitted From Model |  |  | 1.0 | - | - |
|  | Intermediate Risk | 1.790 | 1.420-2.256 | <0.001 |  |  |  |  |  |  | 1.713 | 1.357-2.161 | <0.001 |
|  | High Risk | 2.626 | 1.847-3.734 | <0.001 |  |  |  |  |  |  | 2.409 | 1.687-3.441 | <0.001 |

Three multivariate models were generated. Among other significant factors, Model 1 included the significant individual components of the BSG2020 Guidelines. According to this model, male sex (HR 1.446; p=0.001), left colonic polyps (HR 0.656; p=0.002), multiple index polyps (HR 1.551 for 2-4 polyps and HR 2.340 for 5+ polyps; both p<0.001), and high cytoplasmic SOX9 expression (HR 1.397; p=0.015) were all independent predictive factors of the detection of metachronous polyps or CRC.

Model 2 included BSG2020 Guidelines as its own variable and, omitting its individual components. Here, male sex (HR 1.463; p=0.001), left colonic polyps (HR 0.650; p=0.002), high risk as per the BSG2020 Guidelines (HR 1.677; p<0.001), and high cytoplasmic SOX9 expression (HR 1.465; p=0.005) were all independent predictive factors for metachronous polyp detection.

Model 3 consisted of the combination of cytoplasmic SOX9 with BSG2020 Guidelines, omitting the guidelines components and isolated SOX9 expression. Male sex (HR 1.463; p=0.001), left colonic polyps (0.655; p=0.002), and the “Intermediate Risk” and “High Risk” groups of the combination of SOX9 and BSG2020 Guidelines (HR 1.713 for Intermediate Risk and HR 2.409 for High Risk; p<0.001 for both) were independent predictive factors of metachronous polyps or CRC.

The same analysis was carried out on the test dataset (Table 3), where the combination of BSG2020 criteria and SOX9 protein expression remained independently associated with the clinical outcome.

**Table 3.** Cytoplasmic SOX9 Expression, Clinicopathological Characteristics, and the Detection of Metachronous Polyps or CRC in the INCISE Test Dataset.

|  |  | Univariate |  |  | Multivariate |  |  |  |  |  |  |  |  |
| --- | --- | --- | --- | --- | --- | --- | --- | --- | --- | --- | --- | --- | --- |
|  |  |  |  |  | Model 1: BSG2020 Guidelines Components |  |  | Model 2: BSG2020 Guidelines |  |  | Model 3: Combined Cytoplasmic SOX9 and BSG2020 Guidelines |  |  |
|  |  | Hazard Ratio | 95% CI | P | Hazard Ratio | 95% CI | P | Hazard Ratio | 95% CI | P | Hazard Ratio | 95% CI | P |
| Sex | Female | 1.0 | - | - | - | - | - | - | - | - | - | - | - |
|  | Male | 1.072 | 0.778-1.478 | 0.670 | - | - | - | - | - | - | - | - | - |
| Age | Screening Age | 1.0 | - | - | - | - | - | - | - | - | - | - | - |
|  | Above Screening Age | 1.453 | 0.811-2.603 | 0.209 | - | - | - | - | - | - | - | - | - |
| Site | Right Colon | 1.0 | - | - | 1.0 | - | - | 1.0 | - | - | 1.0 | - | - |
|  | Left Colon | 0.596 | 0.424-0.837 | **0.003** | 0.556 | 0.387-0.798 | **0.001** | 0.595 | 0.416-0.851 | **0.005** | 0.595 | 0.416-0.851 | **0.005** |
|  | Rectum | 0.676 | 0.402-1.137 | 0.140 | 0.544 | 0.313-0.945 | **0.031** | 0.641 | 0.373-1.100 | 0.106 | 0.643 | 0.374-1.105 | 0.110 |
| High Grade Dysplasia | Absent | 1.0 | - | - | - | - | - | Omitted From Model |  |  | Omitted From Model |  |  |
|  | Present | 1.114 | 0.778-1.597 | 0.555 | - | - | - |  |  |  |  |  |  |
| Number of Polyps | 1 | 1.0 | - | - | 1.0 | - | - | Omitted From Model |  |  | Omitted From Model |  |  |
|  | 2-4 | 1.547 | 1.095-2.185 | **0.013** | 1.388 | 0.979-1.967 | 0.066 |  |  |  |  |  |  |
|  | 5+ | 2.584 | 1.711-3.902 | **<0.001** | 2.574 | 1.667-3.974 | **<0.001** |  |  |  |  |  |  |
| Polyp Type | Tubular | 1.0 | - | - | - | - | - | - | - | - | - | - | - |
|  | Tubulovillous | 0.846 | 0.637-1.123 | 0.248 | - | - | - | - | - | - | - | - | - |
|  | Villous | 1.741 | 0.929-3.263 | 0.083 | - | - | - | - | - | - | - | - | - |
| BSG2020 Guidelines | Low Risk | 1.0 | - | - | Omitted From Model |  |  | 1.0 | - | - | Omitted From Model |  |  |
|  | High Risk | 1.687 | 1.218-2.338 | **0.002** |  |  |  | 1.531 | 1.101-2.130 | **0.011** |  |  |  |
| Cytoplasmic SOX9 Adenoma | Low Expression | 1.0 | - | - | 1.0 | - | - | 1.0 | - | - | Omitted From Model |  |  |
|  | High Expression | 1.654 | 1.113-2.457 | **0.013** | 1.548 | 1.039-2.305 | **0.032** | 1.606 | 1.080-2.390 | **0.019** |  |  |  |
| Combined Cytoplasmic SOX9 and BSG2020 Guidelines | Low Risk | 1.0 | - | - | Omitted From Model |  |  | Omitted From Model |  |  | 1.0 | - | - |
|  | Intermediate Risk | 1.614 | 1.129-2.307 | **0.009** |  |  |  |  |  |  | 1.590 | 1.112-2.275 | **0.011** |
|  | High Risk | 2.473 | 1.452-4.209 | **0.001** |  |  |  |  |  |  | 2.413 | 1.413-4.123 | **0.001** |

## DISCUSSION

This study presents a mutational and protein expression analysis of SOX9 in pre-cancerous human colorectal polyps removed as part of the Scottish Bowel Screening Programme. For the first time, it proposes a new method to improve the current BSG2020 Guidelines using adenoma cellular protein expression data.

Demand for colonoscopy services worldwide is increasing[30], raising concerns as to whether these demands can be met. Furthermore, the COVID-19 pandemic exacerbated screening and surveillance backlogs, with a reported drop of 95% of endoscopic services during the height of the pandemic, as reported on the National Endoscopic Database[31]. In addition, endoscopic assessment is not without its complications and as such, improved patient stratification is necessary to mitigate this risk[31]. The current study suggests that high protein expression of SOX9 is potentially beneficial when combined with current clinical parameters to further stratify low and high risk patients, ultimately reducing the demand for endoscopic surveillance.

The current BSG2020 Risk Index consists of polyp(s) number, size, and dysplasia to stratify risk and recommend whether a patient requires a follow-up colonoscopy. It has been suggested that CRC prevention benefits are mainly derived from the initial colonoscopy rather than any subsequent examination[8]. Therefore, it is important to maximise the information derived from the initial colonoscopy. However, when the BSG2020 Guidelines are employed within the INCISE cohort, over a third of the low risk patients develop metachronous polyps or CRC while more than half of the high risk patients do not. The latter are therefore subject to unnecessary procedures which are invasive, risky, and place a burden on the healthcare sector, whilst the former are potentially developing advanced adenomas or cancers without appropriate surveillance. The INCISE collaborative proposes that examination of polyp tissue in greater detail, and using immunohistochemical, genomic, and transcriptomic techniques may improve this risk stratification.

A recent systematic review from our group assessed the current risk stratification methods for metachronous polyp surveillance[15]. This review evaluated studies which examined the use of biomarkers for metachronous polyp risk including mutational signatures in oncogenes and tumour suppressors genes[30,32,33]. Furthermore, it identified genetic abnormalities and protein expression levels including β-catenin[34] as possible predictors of metachronous polyp risk. It concluded that a novel panel of protein markers is required to refine risk stratification adequately.

It is well known that *APC* mutations are important first steps in CRC formation[35,36]. In fact, the involvement of *APC* mutations is early enough that previous work has reported mutations in 85% of tubular polyps and 100% in tubulovillous polyps tested[37]. Likewise, KRAS is mutated in over a third of advanced polyps and CRC[38], with the most cited oncogenic pathways including the MAPK and PI3K pathways. It has been reported that between 40-50% of CRC have *KRAS* mutations[38,39] with many downstream regulators of cell proliferation being affected[40] by genetic changes in the RTK-RAS pathway. This work supports the notions of high mutational burden in early CRC, since *APC* and *KRAS* mutations were the top 2 mutated genes in our samples. Although only the 6^th^ most mutated gene, *SOX9* stood out with its significant co-occurrence with both *APC* and *KRAS*, unlike the higher ranked genes, *MSH3, KMT2C,* and *ZFHX3*. SOX9 is a stemness marker known to interact with the WNT pathway in CRC, either by inhibiting β-catenin, or by directly interacting with TCF transcription factors[41,42], albeit with contradicting evidence with regards to its tumorigenicity. Furthermore, previous work suggests that SOX9 mutations are associated with KRAS mutations[43].

Our results not only point to WNT signaling members as being highly mutated in adenomas (*APC* and *TCF7L2*), and to mutational patterns in WNT direct effectors like *SOX9*, but also to the pathway being affected in most of the samples in the study. The same is true for *RTK-RAS* pathway, where a large portion of our samples have *KRAS* mutations. Although these patterns were found in an exonic sequencing study of CRC patients[44], our study is the first to demonstrate them in colorectal adenomas from a screening population.

We further investigated *SOX9* since it has exhibited both tumor-suppressing and oncogenic effects in the literature[42,45]. These contradicting effects could be attributed to truncating mutations leading to a short isoform of SOX9 protein identified by Abdel-Samad et al.[46]. Regardless, to our knowledge, this is the first study that identified *SOX9* mutations in human adenoma tissue[47].

Previous work suggests that high SOX9 expression is a poor prognostic variable in CRC[48], and is critical for the initiation of CRC[49], however those works do not delve into the significance of the cellular compartment SOX9 is expressed in, with regards to those effects. Here, we show that it is cytoplasmic SOX9 expression (not nuclear[50]) that is significantly associated with metachronous polyps.

Stemness is integral to adenoma-carcinoma progression[51]. High SOX9 expression leading to shorter polyp detection time could reflect the stemness functions of SOX9, which have been shown to block cellular differentiation and progress CRC[47]. Although SOX9 has been reported to have tumor-suppressing functions[52], effects of losing its transactivating domain have been shown to be oncogenic[46]. Here, we demonstrate that stemness markers like SOX9 could be used to improve metachronous polyp prediction if present in the index polyp.

There are several limitations to our current study. The patient cohort was constructed from a single geographical area, therefore despite the use of a local test/validation cohort, external validation of the results is required to account for population heterogeneity. Since this study used TMAs for efficient use of tissue, the results will require validation in full section FFPE adenomas, to both ensure our findings stand against the heterogeneity of tissue and ensure clinical translation as full sections are the used material in clinical practice. Furthermore, the large “Intermediate Risk” group in the final risk score is perhaps a less useful group clinically, and further work will be directed at optimising their stratification. Polyps included were ≥10mm in size, therefore the results need to be investigated smaller polyps common in clinical settings. Finally, this study only covered the protein expression of full length SOX9. A comparison between that and truncated versions known for their oncogenicity[46] is required to fully understand the biological interactions taking place.

In conclusion, this work has demonstrated that current BSG2020 post polypectomy surveillance guidelines, whilst being effective, can be improved upon with the implementation of additional tissue-based assessment. Furthermore, this work suggests that stemness markers like SOX9 could be used to improve risk stratification. This study offers a glimpse of the potential added value of analysis beyond phenotypic histopathological characteristics with the potential to reduce the burden of surveillance endoscopy on patients and in health services.

## Data Availability

Mutational data used in this study are available at Datacite DOI: 10.5525/gla.researchdata.1223, Datacite DOI: 10.5525/gla.researchdata.1307, Datacite DOI: 10.5525/gla.researchdata.1498, and Datacite DOI: 10.5525/gla.researchdata.1602.Data used in this study is available upon reasonable request under condition of collaboration.

