## supplementary for "SOX9 Expression in Colorectal Adenomas Improves Surveillance Colonoscopy Risk Stratification in a Bowel Screening Population"

### SUPPLEMENTARY MATERIALS AND METHODS

#### Tissue Microarray Construction

A Tissue microarray (TMA) was constructed to represent 1257 patients of which 1176 were valid. To account for tissue heterogeneity, two cores were taken from the luminal epithelium (LE), and two from the basal epithelium (BE; S-Fig.1), with a core thickness of 2.5µm.

#### Sample DNA Sequencing Expanded

Sample sequencing and variant calling was performed by the Genomic Innovation Alliance (Glasgow, UK). In brief, 50ng DNA was enzymatically fragmented prior to preparation of sequencing libraries using the Agilent SureSelect XT2 HS2 method (Agilent, Santa Clara, CA). Regions of interest were enriched with the Agilent SureSelect CancerPlus panel (Design ID: S3225252, included genes listed in S-Table 3) and the quality and quantity of libraries determined by TapeStation using a D1000 ScreenTape (5067-5582, Agilent, Santa Clara, CA). The libraries were standarised to 1.5nM for pooling onto 2x 75bp S1v1.5 flow cells (Illumina, San Diego, CA) to run through a NovaSeq 6000. The HOLMES pipeline (v1.3, v1.3.1) was used to process sequencing data and generate files containing single nucleotide variations, structural variations, and copy-number variations. Raw .cblc files from the NovaSeq 6000 were converted to FASTQ files using Bcl2fastq (v2.19.1.403, v2.20.0.422). Alignment was performed with bwa (v0.7.15). Variant calling was carried out using deepSNV/Shearwater (v1.22) for single nucleotide variations, and pindel (v0.2.5b8-ww1) for larger insertions and deletions. Variant annotation was performed using CAVA (v1.2.2.ww1). Brass (v5.3.3-ww10) was used to find structural variation breakpoints by grouping discordant read pairs, and geneCN (v2.1) was used for copy-number calling.

#### Antibody Specificity

##### Selection of Suitable Cell Lines

To check the specificity of our SOX9 antibody, we employed the RStudio DepMap package (Ver. 1.16.0) to identify cell lines that could express our target protein. We identified MCF-7 breast cancer cells as low expressors, and DLD-1 as high expressors (S-Fig.2A-B).

##### Cell Pellet Staining

MCF-7 and DLD-1 cell pellets were kindly provided by Zeanap Mabrouk from the University of Edinburgh. Staining was done by Amna Matly. In brief, following 2 dewaxing steps in Histo-Clear II (HS-202, National Diagnostics, Nottingham, UK) and rehydration in a decreasing alcohol gradient, antigens were retrieved in a heated Tris-EDTA buffer at pH9 under pressure. 3% $H_2O_2$ was used to block exogenous peroxidase activity and 10% casein (2B Scientific, Upper Heyford, UK) was used to block non-specific binding of the antibody. Cell pellets were then stained overnight in 1:500 anti-SOX9 at 4°C. The slides were then washed in TBS-T twice and incubated in ImmPress® (MP-7000, Vector Labs, 2B Scientific, Upper Heyford, UK) for 30 minutes at room temperature, before being washing twice in TBS-T and incubated in ImmPact® DAB chromogen (SK-4105, Vector Labs, 2B Scientific, Upper Heyford, UK) for 5 minutes at room temperature. The cell pellets were then washed in water, counterstained in Hematoxylin Gill III (Cat. 3801540E, Leica Microsystems, Milton Keynes, UK), and dehydrated in a series of increasing alcohols. The stained cell pellets were mounted with Pertex (Cat. SEA-0100-00A, CellPath, Newton, UK). Slides were scanned using Hamamatsu NanoZoomer Digital Scanner and visualized on NDP.serve (3.3.47). Examples of staining are presented in S-Fig.2C-D with negative controls.

#### SDS Gel Electrophoresis and Western Blotting

##### *Cell Lysis*

MCF-7 and DLD-1 cells were seeded at $2.5x10^5$ cells per well in 6-well plates until they reached 100% confluency. They were then washed in ice cold PBS (Ref. 14190-094, Gibco, Altrincham, UK) and scraped with 150µL of preheated Laemmli sample buffer (63mM Tris-HCL, 2mMM $Na_4P_2O_7$, 5mM EDTA, 10% (v/v) glycerol, 2% (w/v) SDS, 50mM DTT, 0.007 (w/v) bromophenol blue). Cells were sheared by aggressive plunging through a 23-guage needle and syringe, and lysates were boiled for 5 minutes.

##### *SDS-PAGE and Western Blotting*

10% SDS polyacrylamide slab gels were used to perform electrophoresis on MCF-7 and DLD-1 cell lysates. Proteins were then blotted on nitrocellulose paper (Product No.: 10600002, Amersham™Protran™, Germany). Non-specific binding was blocked in 3% bovine serum albumin (BSA; Cat. 05482-100G, Sigma, Gillingham, Dorset, UK) in NATT buffer (pH 7.4; 150mM NaCl, 20mM Tris, 0.368mM TWEEN in dH2O) on a shaking platform for 2 hours at RT. Anti-SOX9 was diluted in 0.3% BSA in NATT, and the nitrocellulose membranes were incubated in the antibody overnight at 4°C on a rolling platform. Membranes were washed 4 times in NATT at 20 minute intervals and incubated in anti-rabbit IgG HRP-linked antibody (Cat. 7074S, Cell Signaling, Danvers, MA, USA) at 1:5000 with Precision Protein StrepTactin-HRP Conjugate (Cat. 1610381, BioRad, Watford, Hertfordshire, UK) in 0.3% BSA in NATT for 90 minutes at RT on a shaking platform. The membranes were washed in NATT a further 4 times and developed using the Pierce ECL Western Blotting Substrate kit (Ref. 32106, ThermoScientific, Altrincham, Cheshire, UK), and imaged using the LI-COR Odessey FC Scanner (LI-COR Biosciences, Ltd., Milton, Cambridge, UK). Rb pAb to β-Tubulin HRP (Cat. Ab21058, Abcam, Trumpington, Cambridge, UK) was used as a loading control at a concentration of 1:5000 in 0.3% BSA in NATT with Precision Protein StrepTactin-HRP Conjugate for 90 minutes at RT on a shaking platform. This was done in technical duplicates and biological triplicates. A representative membrane is shown in S-Fig.2E.

*Protein Quantification*

Resulting bands were quantified on ImageJ (Ver. 1.54d, Java 1.8.0_345 64bit, NIH, USA) and were normalized against the loading control. Related statistical analysis and graphs were carried out and generated on GraphPad Prism (Ver. 10.0.3 (275) 64bit, San Diego, CA, USA; S-Fig.2F).

### RNA Sequencing Expanded

TempO-Seq™ (Biospyder Technologies, Carlsbad, CA, USA) whole transcriptome profiling was performed on 816 patients from the INCISE cohort, according to the manufacturer's instructions using whole FFPE tissue sections. Briefly, FFPE tissue was 3paraffinized prior to digestion. Crude tissue lysates were used as input for whole transcriptome analysis using the Human Whole Transcriptome v2.0 panel. Detector oligos, consisting of a sequence complementary to an mRNA target plus a universal primer landing site, were annealed in immediate juxtaposition to each other on the targeted RNA template and ligated[53]. Amplification of ligated oligos was performed using a unique primer set for each sample, introducing a sample-specific barcode and Illumina adaptors. Barcoded samples were pooled into a single library and run on an Illumina HiSeq 2500 High Output v4 flow cell. Sequencing reads were demultiplexed using BCL2FASTQ software (Illumina, USA). FASTQ files were aligned to the Human Whole Transcriptome v2.0 panel, which consists of 22,537 probes, using STAR[54]. Up to two mismatches were allowed in the 50-nucleotide sequencing read.
