## Supplementary material for "SOX9 Expression in Colorectal Adenomas Improves Surveillance Colonoscopy Risk Stratification in a Bowel Screening Population": S-Fig.

**Supplementary Figures**

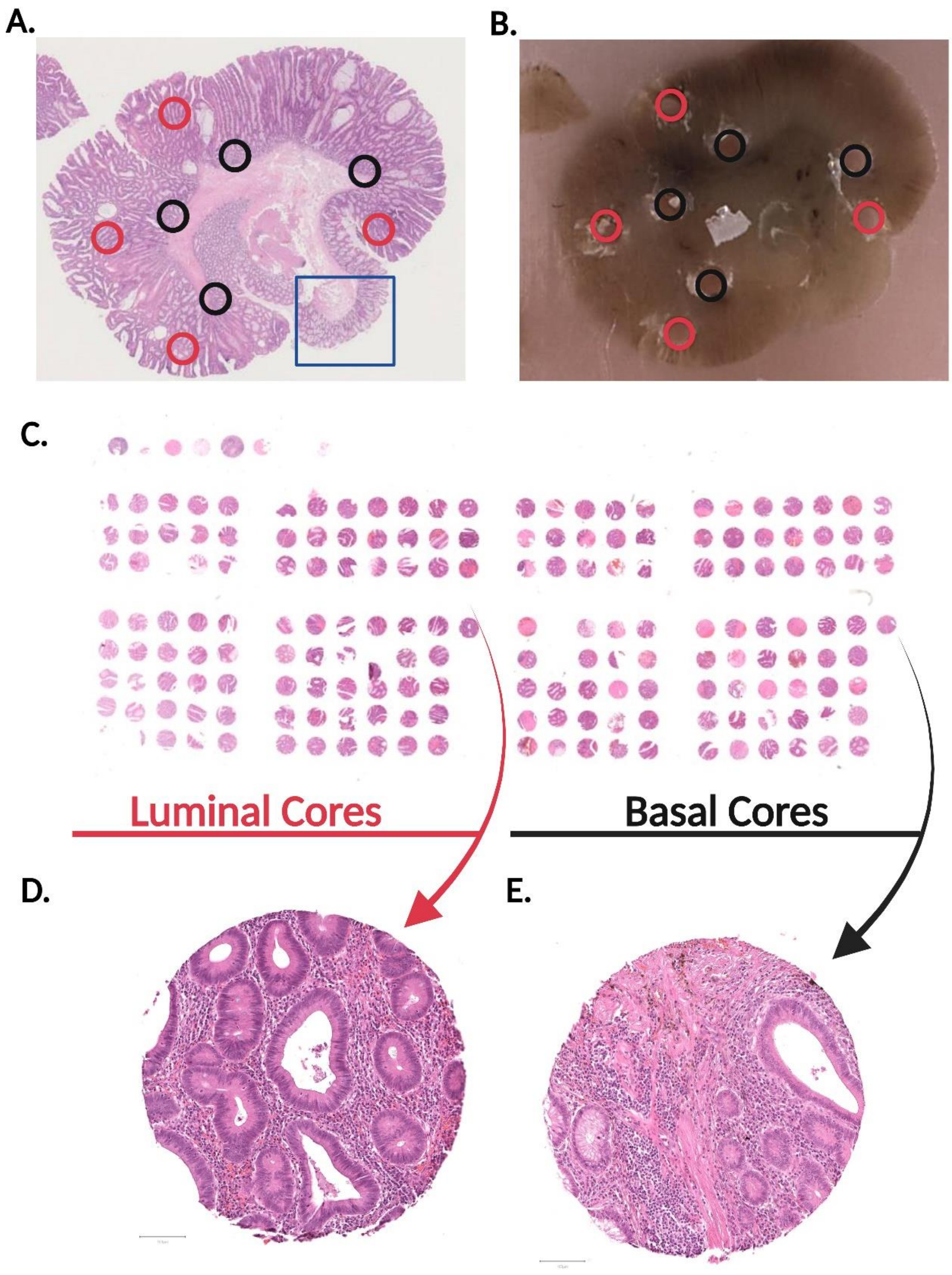


**Supplementary Figure 1. TMA Construction. [A]** Hematoxylin & Eosin (H&E) image of a sample polyp indicating areas designated as luminal (red circles) and basal (black) circle. **[B]** Gross polyp block cored at annotated locations. **[C]** H&E TMA slide where luminal cores are on the left, and basal cores of the same patients at the same location are on the right. **[D]** H&E of luminal polyp core. **[E]** H&E of basal polyp core. D and E are of the same patient.

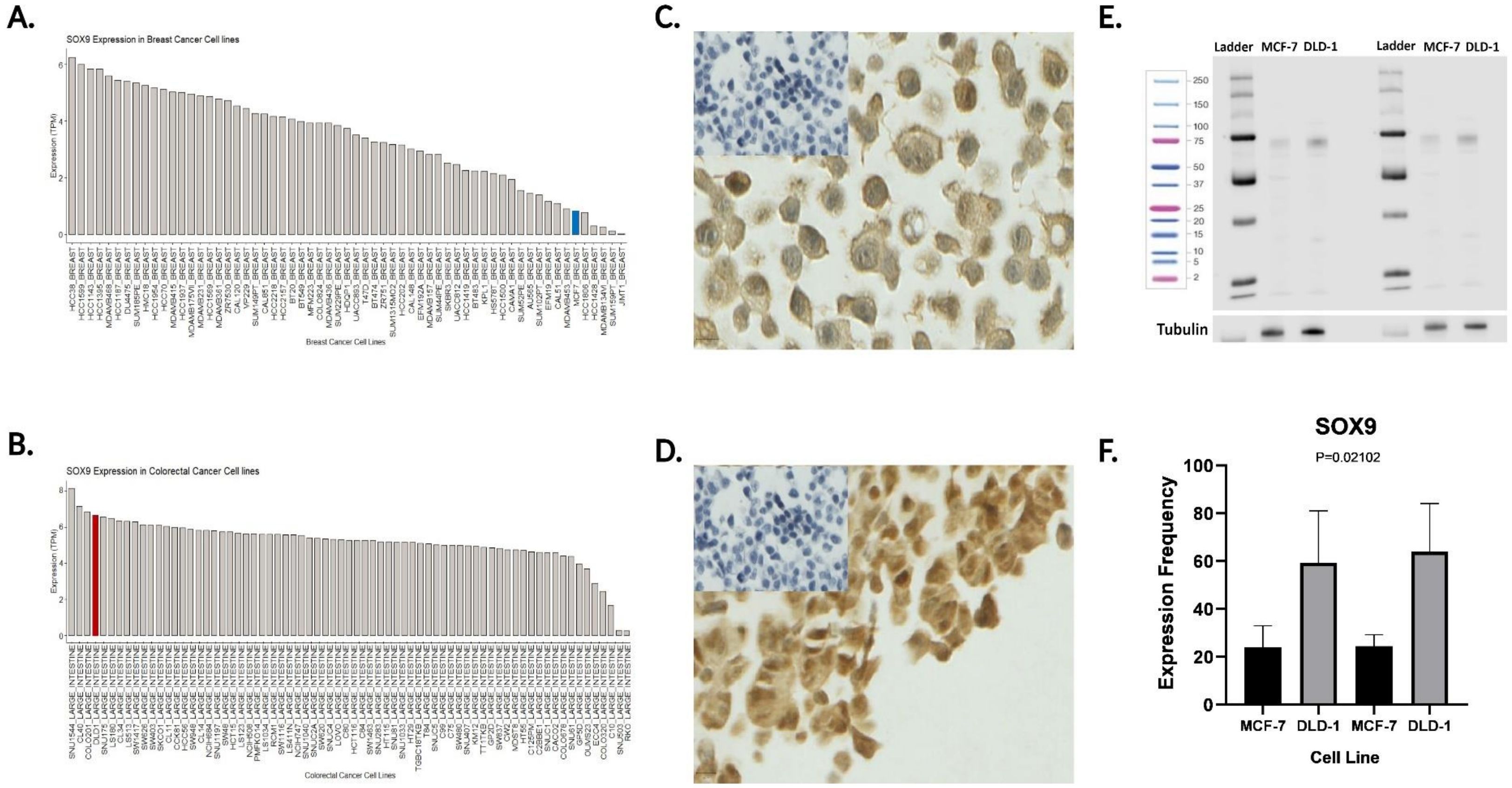


**Supplementary Figure 2. SOX9 Antibody Specificity Check.** DepMap Cell Line Selector of **[A]** breast cancer cell lines, and **[B]** colorectal cancer cell lines that have low (MCF-7, blue bar) and high (DLD-1, red bar) SOX9 expression. Cell pellets of **[C]** MCF-7 breast cancer cell line stained with SOX9, showing lower expression than cell pellets of **[D]** DLD-1 colorectal cancer cell line, with negative no antibody controls in the upper left corners. **[E]** Example western blot of MCF-7 and DLD-1 cell lysates showing varying expression levels as depicted by DepMap and cell pellet staining. β-Tubulin was used as a loading control. Western blots were run in biological triplicates and experimental duplicates. **[F]** Quantification of lysate expression of SOX9, indicating a significant difference between expression levels of MCF-7 cells and DLD-1 cells.

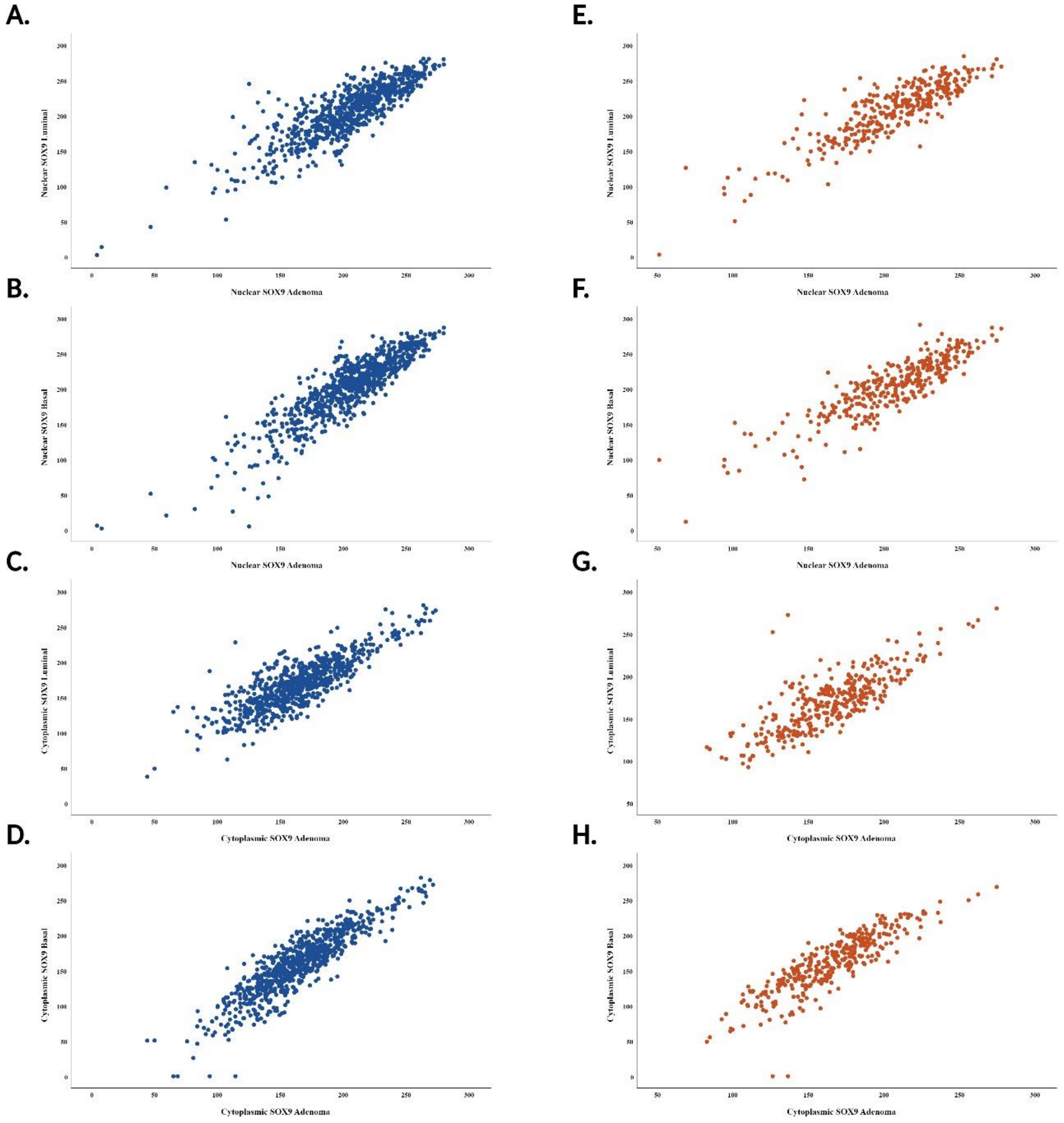


**Supplementary Figure 3. Correlation of Polyp Histological Regions Against Total Polyp Adenomas in the INCISE Training and Test Datasets.** Scatter plot of nuclear SOX9 expression in the luminal epithelium of the polyp vs nuclear SOX9 expression in the total adenoma of the polyp in the **[A]** training and **[E]** test datasets. Scatter plot of nuclear SOX9 expression in the basal epithelium of the polyp vs nuclear SOX9 expression in the total adenoma of the polyp in the **[B]** training and **[F]** test datasets. Scatter plot of cytoplasmic SOX9 expression in the luminal epithelium of the polyp vs cytoplasmic SOX9 expression in the total adenoma of the polyp in the **[C]** training and **[G]** test datasets. Scatter plot of cytoplasmic SOX9 expression in the basal epithelium of the polyp vs cytoplasmic SOX9 expression in the total adenoma of the polyp in the **[D]** training and **[H]** test datasets.

**A.**

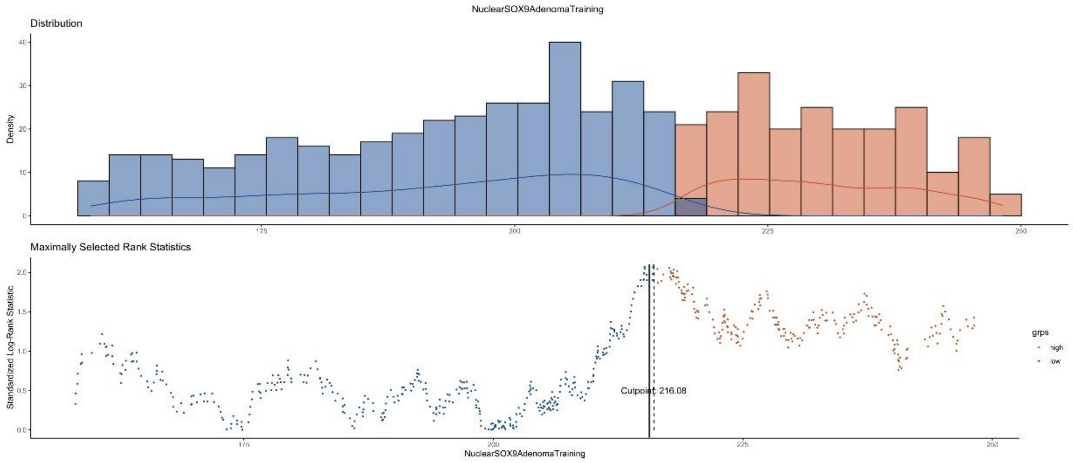


**B.**

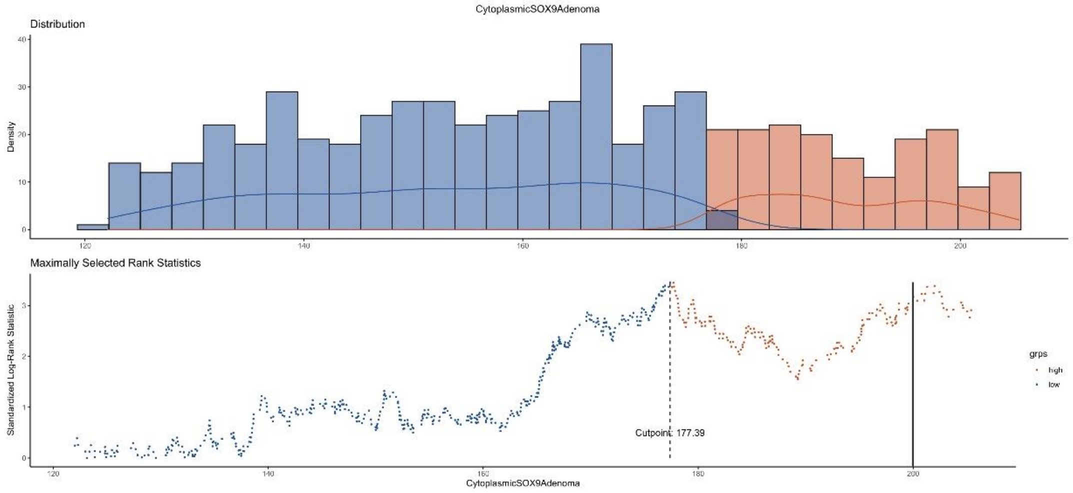


**Supplementary Figure 4. Thresholding of H-Scores. [A]** Threshold generation of nuclear and **[B]** cytoplasmic SOX9 using maximally selected rank statistics on the training dataset. Distribution by density and standardized log rank maximally selected statistic by SOX9 H-Score distribution were both grouped in intervals of 20. The block lines indicate the chosen nuclear threshold of 215 and the cytoplasmic threshold of 200.

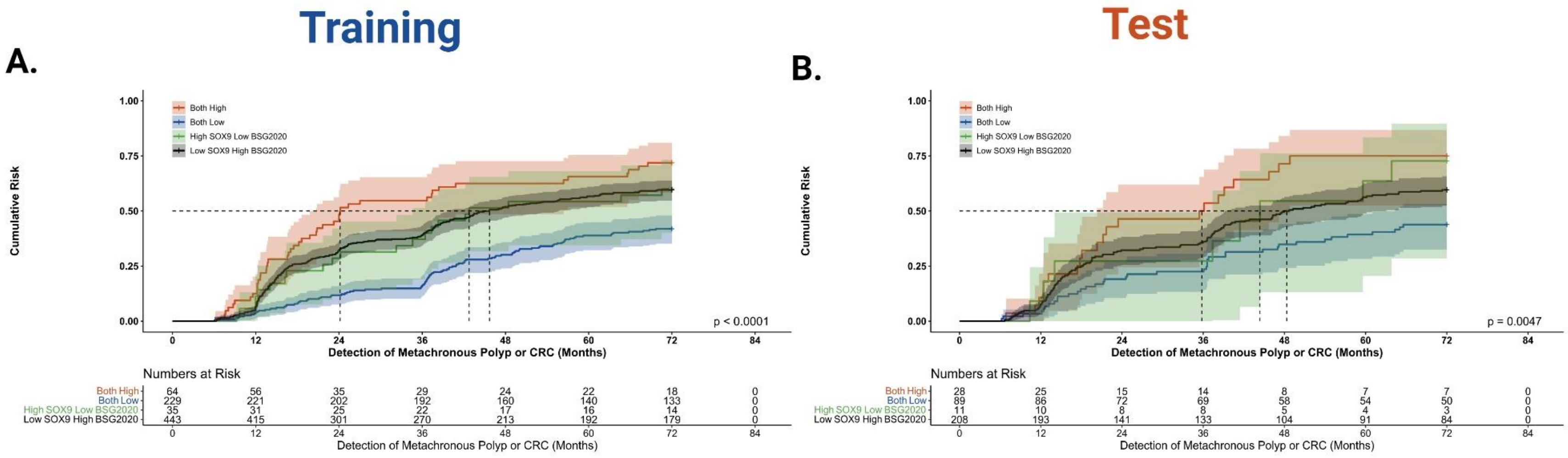


**Supplementary Figure 5. Combination of SOX9 with BSG2020 guidelines.** Patients in the “high for either” groups (panels A and B, black and green groups) were combined into a single risk group “Intermediate Risk” due to large overlap in the final stratification. **[A]** Training dataset. **[B]** Test dataset. Dotted lines are time to median risk (low risk patients do not make it to median risk and so do not have a line).
