## Supplementary material for "SOX9 Expression in Colorectal Adenomas Improves Surveillance Colonoscopy Risk Stratification in a Bowel Screening Population": S-Table

Supplementary Table 1. Cohort Description as Included in the Analysis

| | | Mutational Cohort | | Training Dataset | | Test Dataset | |
|---|---|---|---|---|---|---|---|
| | | **598** | **%** | **818** | **70%** | **358** | **30%** |
| **Sex** | **Female** | 152 | 25 | 234 | 29% | 90 | 25% |
| | **Male** | 446 | 75 | 584 | 71% | 268 | 75% |
| **Age** | **Screening Age** | 570 | 95 | 785 | 96% | 341 | 95% |
| | **Above Screening Age** | 28 | 5 | 33 | 4% | 17 | 5% |
| **Site** | **Right Colon** | 102 | 17 | 100 | 12% | 61 | 17% |
| | **Left Colon** | 421 | 71 | 595 | 73% | 259 | 73% |
| | **Rectum** | 73 | 12 | 121 | 15% | 37 | 10% |
| **High Grade Dysplasia** | **Absent** | 504 | 84 | 686 | 84% | 300 | 84% |
| | **Present** | 94 | 16 | 132 | 16% | 58 | 16% |
| **Number of Polyps** | **1** | 186 | 31 | 278 | 34% | 102 | 29% |
| | **2-4** | 297 | 50 | 440 | 54% | 194 | 54% |
| | **5+** | 115 | 19 | 100 | 12% | 62 | 17% |
| **Polyp Type** | **Tubular** | 239 | 40 | 364 | 45% | 144 | 40% |
| | **Tubulovillous** | 315 | 53 | 396 | 48% | 198 | 55% |
| | **Villous** | 41 | 7 | 57 | 7% | 16 | 5% |
| **BSG2020 Guidelines** | **Low Risk** | 200 | 33 | 280 | 34% | 105 | 29% |
| | **High Risk** | 398 | 67 | 538 | 66% | 253 | 71% |
| **Detection of Metachronous Polyps or CRC** | **No** | 233 | 39 | 363 | 44% | 153 | 43% |
| | **Yes** | 365 | 61 | 455 | 56% | 205 | 57% |

**Supplementary Table 2. Relationship between Cytoplasmic SOX9 Adenoma Expression and Clinicopathological Characteristics in the INCISE Cohort**

| | | Training N=771 | | | | | Test N=336 | | | | |
|---|---|---|---|---|---|---|---|---|---|---|---|
| | | Low Expression | | High Expression | | P | Low Expression | | High Expression | | P |
| | | **672** | **87%** | **99** | **13%** | | **297** | **88%** | **39** | **12%** | |
| **Sex** | **Female** | 196 | 29% | 25 | 25% | 0.421 | 76 | 26% | 8 | 21% | 0.491 |
| | **Male** | 476 | 71% | 74 | 75% | | 221 | 74% | 31 | 79% | |
| **Age** | **50-74 (Screening Age)** | 642 | 96% | 96 | 97% | 0.510 | 287 | 97% | 34 | 87% | **0.007** |
| | **75+ (Above Screening Age)** | 30 | 4% | 3 | 3% | | 10 | 3% | 5 | 13% | |
| **Site** | **Right Colon** | 79 | 12% | 19 | 19% | **<0.001** | 45 | 15% | 9 | 23% | 0.201 |
| | **Left Colon** | 507 | 76% | 49 | 50% | | 222 | 75% | 24 | 62% | |
| | **Rectum** | 85 | 12% | 31 | 31% | | 29 | 10% | 6 | 15% | |
| **High Grade Dysplasia** | **Absent** | 567 | 84% | 78 | 79% | 0.160 | 253 | 85% | 28 | 72% | **0.034** |
| | **Present** | 105 | 16% | 21 | 21% | | 44 | 15% | 11 | 28% | |
| **Number of Polyps** | **1** | 227 | 34% | 35 | 35% | 0.321 | 88 | 30% | 9 | 23% | 0.693 |
| | **2-4** | 368 | 55% | 48 | 49% | | 162 | 54% | 23 | 59% | |
| | **5+** | 77 | 11% | 16 | 16% | | 47 | 16% | 7 | 18% | |
| **Polyp Type** | **Tubular** | 301 | 45% | 41 | 42% | **0.009** | 120 | 40% | 15 | 39% | 0.935 |
| | **Tubulovillous** | 329 | 49% | 45 | 46% | | 165 | 56% | 22 | 56% | |
| | **Villous** | 42 | 6% | 12 | 12% | | 12 | 4% | 2 | 5% | |
| **BSG2020 Guidelines** | **Low Risk** | 229 | 34% | 35 | 35% | 0.803 | 89 | 30% | 11 | 28% | 0.821 |
| | **High Risk** | 443 | 66% | 64 | 65% | | 208 | 70% | 28 | 72% | |
| **Metachronous Polyps or CRC Detection** | **No** | 312 | 46% | 32 | 32% | **0.008** | 134 | 45% | 10 | 26% | **0.021** |
| | **Yes** | 360 | 54% | 67 | 68% | | 163 | 55% | 29 | 74% | |

Supplementary Table 3. List of Genes in Agilent SureSelect CancerPlus Panel

| Gene | Number of Patients Affected by Mutations | Rank |
|---|---|---|
| ABL1 | 15 | 129 |
| ABL2 | 14 | 139 |
| ABR | 9 | 201 |
| ABRAXAS1 | 4 | 281 |
| ACVR1B | 8 | 212 |
| ACVR2A | 34 | 45 |
| AJUBA | 5 | 258 |
| AKAP9 | 44 | 28 |
| AKT1 | 4 | 282 |
| AKT2 | 1 | 320 |
| AKT3 | 5 | 259 |
| ALK | 27 | 75 |
| ALOX12B | 10 | 184 |
| ALOX15B | 13 | 144 |
| AMER1 | 42 | 32 |
| APC | 475 | 1 |
| APLNR | 5 | 260 |
| AR | 25 | 84 |
| ARAF | 14 | 141 |
| ARHGAP35 | 30 | 58 |
| ARID1A | 77 | 9 |
| ARID1B | 40 | 34 |
| ARID2 | 49 | 20 |
| ARID5B | 17 | 121 |
| ASXL1 | 37 | 40 |
| ASXL2 | 31 | 54 |
| ATM | 48 | 22 |
| ATR | 34 | 46 |
| ATRX | 49 | 21 |
| AURKA | 0 | 331 |
| AURKB | 5 | 257 |
| AURKC | 9 | 202 |
| AXIN1 | 7 | 231 |
| AXIN2 | 31 | 55 |
| AXL | 0 | 332 |
| B2M | 1 | 321 |
| BAP1 | 13 | 145 |
| BARD1 | 15 | 131 |
| BCL2 | 0 | 333 |
| BCOR | 52 | 17 |
| BIRC3 | 6 | 243 |
| BLM | 26 | 79 |
| BRAF | 31 | 56 |
| BRCA1 | 28 | 70 |
| BRCA2 | 56 | 16 |
| BRIP1 | 18 | 113 |
| CARD11 | 21 | 97 |
| CASP8 | 14 | 142 |

| | | |
|---|---|---|
| CBFB | 4 | 283 |
| CBL | 15 | 133 |
| CCND1 | 2 | 309 |
| CCND2 | 0 | 334 |
| CCND3 | 5 | 261 |
| CCNE1 | 5 | 262 |
| CD274 | 10 | 185 |
| CD58 | 3 | 293 |
| CD74 | 2 | 310 |
| CDC73 | 11 | 164 |
| CDH1 | 17 | 122 |
| CDK12 | 18 | 115 |
| CDK2 | 2 | 311 |
| CDK4 | 3 | 294 |
| CDK6 | 1 | 322 |
| CDK8 | 13 | 150 |
| CDKN1A | 2 | 312 |
| CDKN1B | 4 | 279 |
| CDKN1C | 1 | 323 |
| CDKN2A | 2 | 313 |
| CDKN2B | 5 | 263 |
| CDKN2C | 3 | 295 |
| CHD4 | 46 | 24 |
| CHD8 | 38 | 39 |
| CHEK1 | 5 | 264 |
| CHEK2 | 11 | 169 |
| CIC | 17 | 123 |
| CIITA | 23 | 91 |
| CKS1B | 1 | 324 |
| COL17A1 | 23 | 87 |
| CREBBP | 32 | 50 |
| CTCF | 11 | 165 |
| CTLA4 | 4 | 284 |
| CTNNB1 | 39 | 37 |
| CUL3 | 45 | 27 |
| CUX1 | 26 | 77 |
| CYLD | 8 | 213 |
| DAXX | 7 | 229 |
| DDR2 | 19 | 109 |
| DDX3X | 5 | 265 |
| DDX5 | 7 | 232 |
| DEFB134 | 0 | 335 |
| DHX9 | 25 | 82 |
| DICER1 | 30 | 61 |
| DNMT1 | 13 | 146 |
| DNMT3A | 6 | 244 |
| EGFR | 20 | 104 |
| EIF4A2 | 3 | 296 |
| ELF3 | 36 | 42 |
| ELOC | 3 | 297 |

| | | |
|---|---|---|
| EP300 | 38 | 38 |
| EPHA2 | 10 | 186 |
| EPHA3 | 20 | 105 |
| ERBB2 | 20 | 102 |
| ERBB3 | 28 | 71 |
| ERBB4 | 18 | 116 |
| ERCC2 | 13 | 147 |
| ERCC3 | 17 | 119 |
| ERCC4 | 33 | 49 |
| ERCC5 | 0 | 336 |
| ERG | 9 | 203 |
| ESR1 | 17 | 124 |
| ETV1 | 10 | 187 |
| ETV4 | 5 | 266 |
| ETV5 | 42 | 33 |
| ETV6 | 11 | 170 |
| EZH2 | 8 | 214 |
| FANCA | 29 | 66 |
| FANCC | 5 | 267 |
| FANCD2 | 23 | 88 |
| FANCE | 6 | 245 |
| FANCF | 6 | 241 |
| FANCG | 11 | 166 |
| FANCL | 3 | 298 |
| FANCM | 65 | 13 |
| FAS | 4 | 285 |
| FAT1 | 74 | 11 |
| FBXW7 | 75 | 10 |
| FGF19 | 1 | 325 |
| FGFR1 | 6 | 246 |
| FGFR2 | 10 | 188 |
| FGFR3 | 11 | 171 |
| FGFR4 | 10 | 189 |
| FLT1 | 18 | 114 |
| FOXA1 | 3 | 299 |
| FOXA2 | 6 | 247 |
| FOXL2 | 2 | 314 |
| FOXO1 | 8 | 215 |
| FOXP1 | 0 | 337 |
| FUBP1 | 10 | 181 |
| GATA3 | 11 | 172 |
| GATA6 | 21 | 99 |
| GNA11 | 3 | 300 |
| GNA13 | 2 | 315 |
| GNAQ | 1 | 326 |
| GNAS | 33 | 48 |
| GPS2 | 9 | 204 |
| H3F3A | 0 | 338 |
| H3F3B | 0 | 339 |
| HGF | 4 | 280 |

| | | |
|---|---|---|
| HIF1A | 11 | 167 |
| HIST1H1C | 0 | 340 |
| HIST1H3B | 0 | 341 |
| HIST1H3C | 0 | 342 |
| HIST2H3C | 0 | 343 |
| HLA-A | 64 | 14 |
| HLA-B | 23 | 86 |
| HLA-C | 20 | 106 |
| HNF1A | 11 | 168 |
| HRAS | 5 | 268 |
| IDH1 | 6 | 248 |
| IDH2 | 6 | 249 |
| IDO1 | 6 | 250 |
| IDO2 | 13 | 151 |
| IFNGR1 | 7 | 233 |
| IFNGR2 | 8 | 216 |
| IGF1R | 0 | 344 |
| IL6ST | 9 | 205 |
| IRF1 | 0 | 345 |
| JAK1 | 17 | 125 |
| JAK2 | 10 | 182 |
| JAK3 | 5 | 269 |
| JUN | 5 | 270 |
| KDM5C | 15 | 134 |
| KDM6A | 15 | 130 |
| KDR | 24 | 85 |
| KEAP1 | 10 | 190 |
| KIT | 15 | 135 |
| KLF4 | 7 | 234 |
| KMT2A | 34 | 44 |
| KMT2B | 31 | 57 |
| KMT2C | 97 | 4 |
| KMT2D | 79 | 8 |
| KRAS | 189 | 2 |
| LZTR1 | 13 | 148 |
| MAP2K1 | 8 | 217 |
| MAP2K2 | 5 | 271 |
| MAP2K4 | 9 | 206 |
| MAP3K1 | 29 | 63 |
| MAPK1 | 7 | 230 |
| MAX | 1 | 327 |
| MCL1 | 7 | 235 |
| MDM2 | 3 | 301 |
| MDM4 | 9 | 199 |
| MECOM | 10 | 191 |
| MED12 | 27 | 76 |
| MEN1 | 4 | 286 |
| MET | 18 | 112 |
| MGA | 80 | 7 |
| MGMT | 5 | 272 |

| | | |
|---|---|---|
| MLH1 | 15 | 136 |
| MRE11 | 10 | 192 |
| MSH2 | 21 | 100 |
| MSH3 | 25 | 80 |
| MSH6 | 28 | 68 |
| MTOR | 22 | 92 |
| MUTYH | 10 | 193 |
| MYB | 7 | 236 |
| MYC | 4 | 287 |
| MYCL | 5 | 273 |
| MYCN | 5 | 274 |
| MYH9 | 20 | 103 |
| NAB2 | 6 | 251 |
| NBN | 12 | 153 |
| NCOA2 | 32 | 51 |
| NCOR1 | 28 | 69 |
| NF1 | 50 | 19 |
| NF2 | 12 | 154 |
| NFE2L2 | 9 | 198 |
| NLRC5 | 28 | 72 |
| NOTCH1 | 29 | 64 |
| NOTCH2 | 35 | 43 |
| NOTCH3 | 30 | 59 |
| NOTCH4 | 44 | 29 |
| NPM1 | 6 | 252 |
| NRAS | 28 | 73 |
| NRG1 | 22 | 94 |
| NSD1 | 40 | 35 |
| NSD3 | 25 | 81 |
| NTRK1 | 12 | 155 |
| NTRK2 | 16 | 128 |
| NTRK3 | 19 | 110 |
| PALB2 | 29 | 65 |
| PARP1 | 23 | 89 |
| PAX5 | 12 | 156 |
| PBRM1 | 22 | 95 |
| PCBP1 | 45 | 26 |
| PDCD1LG2 | 2 | 316 |
| PDGFRA | 0 | 346 |
| PDGFRB | 12 | 157 |
| PHF6 | 3 | 302 |
| PIAS3 | 11 | 173 |
| PIAS4 | 6 | 253 |
| PIK3CA | 32 | 52 |
| PIK3CB | 7 | 237 |
| PIK3CD | 8 | 218 |
| PIK3R1 | 23 | 90 |
| PIK3R2 | 6 | 242 |
| PIM1 | 3 | 303 |
| PLCG1 | 15 | 137 |

| | | |
|---|---|---|
| PMS1 | 11 | 174 |
| PMS2 | 21 | 98 |
| POLE | 52 | 18 |
| POLQ | 45 | 25 |
| PPM1D | 10 | 180 |
| PPP2R1A | 3 | 304 |
| PPP2R2A | 1 | 328 |
| PPP4R2 | 6 | 254 |
| PPP6C | 2 | 308 |
| PRKAR1A | 10 | 194 |
| PSIP1 | 13 | 149 |
| PTCH1 | 21 | 101 |
| PTEN | 5 | 275 |
| PTK2 | 9 | 207 |
| PTPN11 | 12 | 158 |
| PTPRD | 36 | 41 |
| QSER1 | 22 | 93 |
| RAC1 | 2 | 317 |
| RAD21 | 6 | 255 |
| RAD50 | 0 | 347 |
| RAD51B | 8 | 219 |
| RAD51C | 7 | 238 |
| RAD51D | 0 | 348 |
| RAD52 | 8 | 220 |
| RAD54L | 8 | 221 |
| RAF1 | 11 | 175 |
| RASA1 | 16 | 126 |
| RB1 | 12 | 159 |
| RBM10 | 30 | 60 |
| RET | 15 | 138 |
| RFX5 | 12 | 160 |
| RFXAP | 4 | 288 |
| RHEB | 1 | 329 |
| RHOA | 4 | 289 |
| RICTOR | 18 | 117 |
| RIT1 | 0 | 349 |
| RNF43 | 11 | 163 |
| ROS1 | 27 | 74 |
| RPL22 | 12 | 161 |
| RPL5 | 8 | 222 |
| RUNX1 | 11 | 176 |
| SERPINB3 | 8 | 209 |
| SERPINB4 | 7 | 239 |
| SETBP1 | 19 | 108 |
| SETD2 | 42 | 31 |
| SF3B1 | 20 | 107 |
| SLC34A2 | 17 | 118 |
| SMAD2 | 5 | 276 |
| SMAD3 | 2 | 318 |
| SMAD4 | 26 | 78 |

| | | |
|---|---|---|
| SMARCA4 | 29 | 62 |
| SMARCB1 | 12 | 162 |
| SMC1A | 13 | 152 |
| SMC3 | 8 | 210 |
| SMG1 | 57 | 15 |
| SMO | 16 | 127 |
| SOCS1 | 3 | 305 |
| SOS1 | 17 | 120 |
| SOX17 | 11 | 177 |
| SOX9 | 90 | 5 |
| SPEN | 47 | 23 |
| SPOP | 2 | 319 |
| SRC | 5 | 277 |
| SRSF2 | 1 | 330 |
| STAG1 | 14 | 143 |
| STAG2 | 6 | 256 |
| STAT1 | 9 | 208 |
| STAT3 | 8 | 211 |
| STAT5B | 10 | 195 |
| STK11 | 4 | 290 |
| SYK | 10 | 196 |
| TAF1 | 29 | 67 |
| TAF3 | 14 | 140 |
| TAP1 | 9 | 200 |
| TAP2 | 5 | 278 |
| TAPBP | 3 | 306 |
| TBL1XR1 | 8 | 223 |
| TBX3 | 15 | 132 |
| TCF12 | 11 | 178 |
| TCF7L2 | 66 | 12 |
| TERT | 0 | 350 |
| TET2 | 34 | 47 |
| TGFBR1 | 32 | 53 |
| TGFBR2 | 7 | 240 |
| TMPRSS2 | 3 | 307 |
| TP53 | 81 | 6 |
| TP53BP1 | 39 | 36 |
| TP73 | 8 | 224 |
| TRAF7 | 8 | 225 |
| TSC1 | 19 | 111 |
| TSC2 | 43 | 30 |
| U2AF1 | 0 | 351 |
| UVRAG | 8 | 226 |
| VHL | 3 | 292 |
| WRN | 22 | 96 |
| WT1 | 4 | 291 |
| XBP1 | 7 | 228 |
| XPO1 | 11 | 179 |
| YAP1 | 0 | 352 |
| ZFHX3 | 99 | 3 |

| | | |
|---|---|---|
| ZFP36L1 | 10 | 197 |
| ZMYM2 | 25 | 83 |
| ZMYM3 | 8 | 227 |
| ZNF703 | 10 | 183 |
| ZNF750 | 0 | 353 |

**Supplementary Table 4. Top 10 Somatic Interactions for Mutated Gene Pairs**

| Rank | Gene 1 | Gene 2 | Event | Adjusted P-Value | Statistically Significant | Event Ratio (Gene 1/Gene2) | Event Ratio (%) |
|---|---|---|---|---|---|---|---|
| 1 | *TCF7L2* | *KRAS* | Co-occurrence | 0.009285972 | Yes | 45/203 | 22 |
| 2 | *MGA* | *KMT2C* | Co-occurrence | 0.010712746 | Yes | 28/144 | 19 |
| 3 | *SOX9* | *KRAS* | Co-occurrence | 0.010712746 | Yes | 48/206 | 23 |
| 4 | *SOX9* | *APC* | Co-occurrence | 0.046736556 | Yes | 96/432 | 22 |
| 5 | *MGA* | *KMT2D* | Co-occurrence | 0.068406272 | No | 21/134 | 16 |
| 6 | *KRAS* | *APC* | Co-occurrence | 0.082777185 | No | 185/354 | 52 |
| 7 | *ZFHX3* | *KMT2D* | Co-occurrence | 0.094753799 | No | 23/146 | 16 |
| 8 | *MGA* | *APC* | Co-occurrence | 0.320018639 | No | 82/447 | 18 |
| 9 | *MGA* | *SOX9* | Co-occurrence | 0.399320412 | No | 20/149 | 13 |
| 10 | *MSH3* | *KRAS* | Mutually Exclusive | 0.399320412 | No | 35/257 | 14 |

**Supplementary Table 5. Incidence of Metachronous Lesion Types with Combined Cytoplasmic SOX9 and BSG2020 Guidelines**

| | Training | | | | | | | Test | | | | | | |
|---|---|---|---|---|---|---|---|---|---|---|---|---|---|---|
| | **Low Risk** | | **Intermediate Risk** | | **High Risk** | | **P** | **Low Risk** | | **Intermediate Risk** | | **High Risk** | | **P** |
| | **229** | **30** | **47** | **62%** | **64** | **8%** | | **89** | **26%** | **219** | **65%** | **28** | **8%** | |
| **None** | 133 | 58% | 191 | 40% | 18 | 28% | **<0.0001** | 50 | 56% | 87 | 40% | 7 | 25% | **0.014** |
| **Non-Advanced Adenoma** | 66 | 29% | 205 | 43% | 22 | 34% | | 28 | 32% | 97 | 44% | 13 | 46% | |
| **Advanced Adenoma or CRD** | 30 | 13% | 82 | 17% | 24 | 38% | | 11 | 12% | 35 | 16% | 8 | 29% | |
